# Clinical characteristic and imaging findings of post-infarction left ventricular pseudoaneurysm versus aneurysm: a pooled analysis of 21,472 patients

**DOI:** 10.1101/2023.02.23.23286381

**Authors:** Elmira Jafari Afshar, Amirhossein Tayebi, Parham Samimisedeh, Vahid Shahnavaz, Aryan Madady, Hadith Rastad, Neda Shafiabadi Hassani

**Affiliations:** Cardiovascular Research Center, Alborz University of Medical Sciences, Karaj, Iran; Cardiovascular surgery department, Shahid Rajaee hospital, Alborz University of Medical Sciences, Karaj, Iran; Non-communicable Diseases Research Center, Alborz University of Medical Sciences, Karaj, Iran; Herrington Heart and Vascular Institute, University Hospitals Cleveland Medical Center, Cleveland, Ohio, USA

**Author notes:** Hadith Rastad and Neda Shafiabadi Hassani equally contributed as corresponding authors. Elmira Jafari Afshar and Amir Hossein Tayebi equally contributed as first authors.

**Keywords:** Left ventricular pseudoaneurysm, Left ventricular aneurysm, Myocardial infarction, imaging findings, clinical presentations

## Abstract

**Background:** Left ventricular pseudoaneurysm (LVPA) is a rare but life-threatening complication of myocardial infarction (MI). Given the similarities in the clinical presentations and the appearance of the cardiac imaging, differentiation of LVPA from left ventricular aneurysm (LVA) remains a challenge but is imperative for timely management. We summarized and compared clinical and imaging findings of post-MI LVPA and LVA.

**Method:** We performed a comprehensive search of the literature in PubMed and Scopus databases using combinations of key terms covering LVPA / LVA and MI. In both LVA and LVPA, individual-level patient data (IPD) and aggregated-level data (AD) studies were combined through a two-stage analysis method.

**Results:** We identified 379 eligible articles on LVPA (N= 504 patients) and 120 on LVA (n= 20,968). Based on our pooled analysis, cases were predominantly male in both groups (70.4%and 75.7 %, respectively), but LVPA patients were roughly older (Mean (95% Confidence interval (CI): (65.4 (62.4, 68.4) vs. 60.8 (58.9, 62.8) years, respectively) and had a shorter mean time interval from MI to diagnosis than LVA (5.1 vs. 27.8, months). At presentation, while 33.8 (95% CI: 22.1, 46.0) of patients with LVA had arrhythmia, only 1.0 % (95% CI: 0.0, 2.9) of LVPA patients presented with this symptom. LVPA compared to the LVA group, more frequently had ST-segment elevation (43.2% Vs. 28.6, respectively) but less frequently ECG signs of the old MI (42.2% Vs. 61.9, respectively). Echocardiography showed a lower diagnostic value in LVPA than LVA (Sensitivity: 81.4% Vs. 97.5%). Contrary to LVA, LVPA is mainly located on posterior and inferior segments based on echocardiography evaluations. On Cardiac MRI, the majority of LVPA patients had pericardial LGE (84.0% (CI 95%: 63.9, 95.5)). A higher percentage of LVPA compared to the LVA group dead during hospitalization (13.8% vs. 4.7%, respectively) or after discharge (17.5%vs. vs. 9.0%, respectively).

**Conclusion:** Arrhythmia is likely common in LVA patients at presentation but not in LVPA. LVPA is mainly located on the posterior and inferior, and LVA is on the anterior and apical segments. On cardiac MRI, pericardial LGE may suggest the presence of LVPA rather than LVA in suspected patients.

## Introduction

In patients with myocardial infarction (MI), during the acute phase or later, the left ventricular wall may become vulnerable to the formation of aneurysms. Most aneurysms, called true left ventricular aneurysm (LVA), are formed by an expansion of the thinned myocardium and contain residual elements of the myocardial wall. In rare cases, they cause by the localized rupture of an infarcted area, known as false left ventricular aneurysm (LVPA).

As the wall of LVPA is made up of thrombus and/or pericardium without myocardial fibers (Figure.1), it is a more life-threatening entity, yet less known, than LVA (1–3). Still, differentiation between these outpouchings remains challenging and is of great importance for proper management; while LVA is often treated by elective surgery or medical treatment, LVPA mainly requires urgent surgical intervention (4, 5).

**Figure 1:**
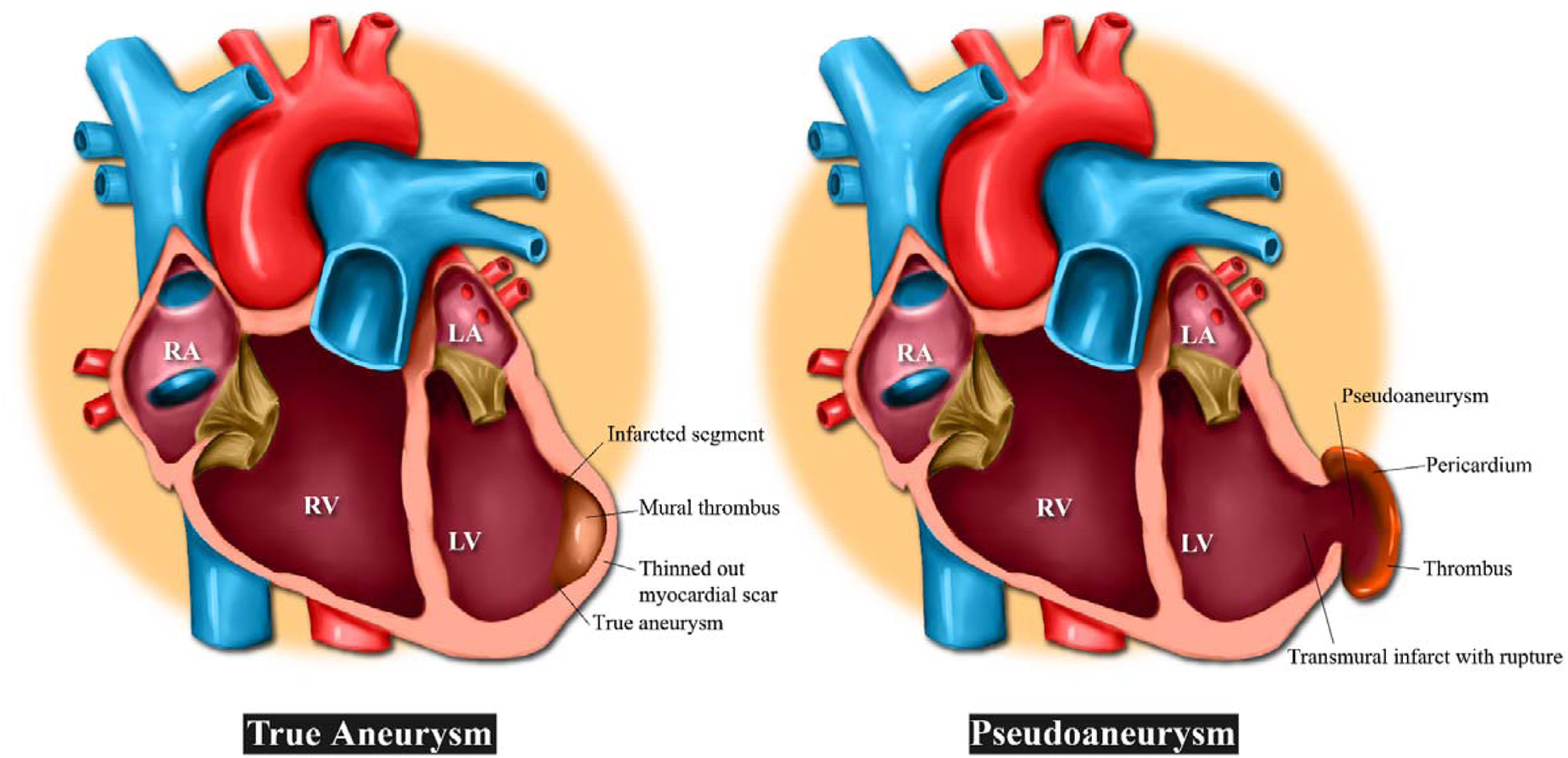
Pseudoaneurysm versus True aneurysm of the left ventricle

While early diagnosis of LVPA and timely management are essential in reducing its mortality rate (6), our understanding of its clinical profile and imaging features is still limited and mainly comes from published case reports or case series. Also, up to our knowledge, no previous review study has compared clinical characteristics and imaging features between LVPA and LVA. In this comprehensive systematic review study, we have pooled data from post-MI cases reported since 1991 to provide better understanding of clinical and imaging features of LVPA. We have also compared these clinical and imaging findings between our pooled cohorts of LVPA and LVA cases to help physicians to distinguish these entities from each other.

## Methods and materials

Our study was conducted according to the Preferred Reporting Items for systematic reviews and Meta-analyses (PRISMA) guidelines (7). As a systematic review and meta-analysis of available literature, ethics committee approval was not required.

We included all eligible observational studies which reported imaging and clinical findings in patients with LVPA after myocardial infarction (MI). To compare these findings with those of the LVA, we also identify all studies with representative samples of patients with LVA following MI.

### Search strategy

We systematically searched online databases, including PubMed and Scopus, with additional manual search in google scholar on September 1st, 2022 using the combination of relevant keywords in two domains:

1. False [and True] Ventricular aneurysm
2. Myocardial infarction

Keywords from each domain were combined using the Boolean operator “OR”, and domains were connected with the “AND” Boolean operator.

Furthermore, we also manually screened the reference list of included and similar articles for possible additional citations. All citations retrieved from databases were imported into EndNote X9 software (version EndNote X9.3.2, Captivate Analytics, California USA), then duplicates were removed.

We presented the detailed search string of each database in Supplementary Tables 1 & 2.

### Study inclusion

The title, abstract, and full text of imported studies were screened independently by two of our researchers [Ej& PS]. Eligible studies that met our inclusion criteria were included for review and meta-analysis. A third senior researcher [HR or NSH] resolved any disagreement between the researchers.

### Selection Criteria

Studies which met all of the following eligibility criteria were included in our review study:

1. Written in English.
2. Included patients with post-MI LVPA [or LVA for the comparison group].
3. Observational study design including case repot, case series, or cohort studies

### Data extraction

Four researchers extracted the data from full-text articles into a data extraction form in Microsoft Excel. (Version 2016, Microsoft Corp., Redmond, WA, USA). We extracted the following data: First author’s name, country, year, study design, sample size, age, sex, types of symptoms, time interval between last MI and diagnosis of the false [and true] aneurysm, cardiovascular disease risk factors and comorbidities, Electrocardiography (ECG), imaging findings [including chest X-ray, echocardiogram, CT-scan, and/or cardiac magnetic resonance imaging (CMR)], treatment strategies, follow-up duration and clinical outcomes, were also extracted from the included studies.

### Risk of bias assessment

The quality of included studies was assessed independently by two of our trained researchers. Any disagreements were resolved with discussion. We used the Joanna Briggs Institute (JBI) critical appraisal tool to appraise the quality of included studies. JBI’s critical appraisal tool contained ten items and its total score ranges from 0 to 10 (8).

### Statistical analyses

Data extracted from case reports and case series with individual-level patient data (IPD) were combined into a single case series. Then this case series was pooled with other studies with aggregated level data (AD) using a standard meta-analysis method. We pooled the categorical variables, including gender, symptoms, risk factors, history of previous surgery, treatment strategy, in-hospital and follow-up death, infarction site, ECGs (electrocardiogram) results, and imaging results using meta-analysis of single proportions; continuous variables such as age, time from myocardial infarction to diagnosis, follow-up duration, and dimension size were combined using meta-analysis of mean and standard deviation. In all meta-analyses, a random-effect model was used to estimate the overall pooled proportion/mean and their 95% confidence intervals (CIs). Meta-analysis was conducted with R-studio version 4.2.2 software.

## Results

Detailed study selection process in detail for LVPA and LVA is presented in Supplementary figure 1 and 2, respectively. After removing duplicates and ineligible articles, we included 379 eligible articles on post-MI LVPA (N= 504 patients) (9–392) and 120 on post-MI LVA (n=20,968). (31,97,145,148,152,158,393–472) Supplementary Table 3. shows the quality of the included case series; all studies scored eight or higher.

### Pseudoaneurysm

Characteristics of each included article are presented in Supplementary Table 4. Regarding the study type, 366 were case reports and 13 case series were published between 1991 to 2022. Cases were reported from 50 countries across the world; over 60% of them were reported from the USA (n=105), Turkey (n=63), and Italy (n=56). Echocardiography, mainly TTE (transthoracic echocardiography), was the most frequently utilized imaging modality for diagnosis. Over 90% (464/504) of reported cases underwent surgical treatment.

Table 1. shows the demographic and clinical characteristics of reported cases. Based on our pooled analysis, patients were predominantly male [70.4 % (95% CI: 65.8, 74.6)] with a mean age of 65.4 years (95% CI: 62.4, 68.4). They most frequently presented with CHF symptoms [45.5% (95% CI: 24.7, 67.1, available data: n=389], followed by dyspnea [43.3 % (95% CI: 24.4, 67.1, n=357], chest pain [39.2%, (95% CI:24.0, 55.4, n=409]; with a less frequency, cardiogenic shock [10.0%, (95% CI: 0.4,58.8, n=352)], syncope [6.0%, (95% CI: 3.5, 8.9, n=340], and arrhythmia [1.0% (95% CI:0.0, 2.9, n=241)] were also presented in these patients.

**Table 1.**
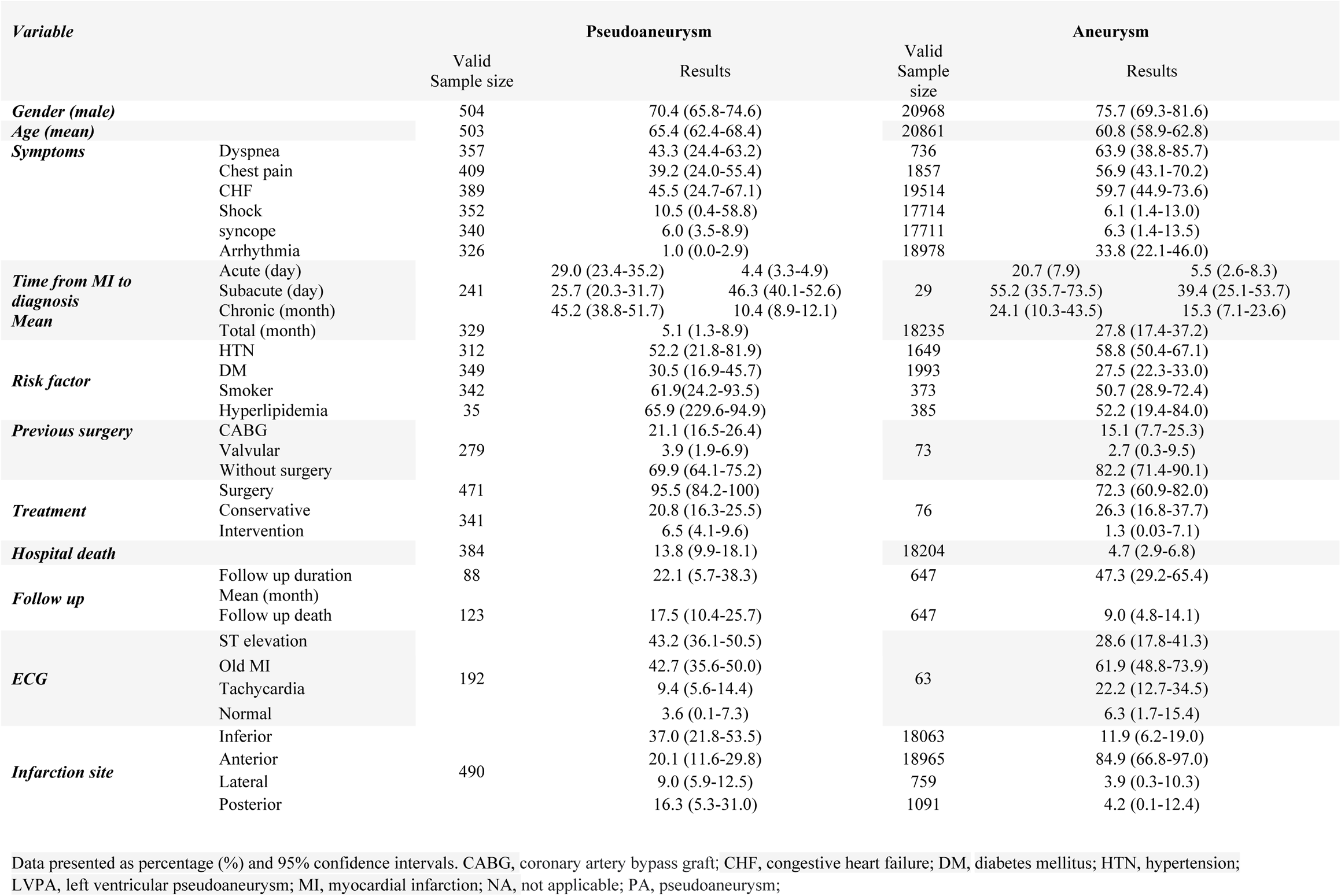
Clinical characteristics of LVPA and LVA patients

The time interval between the last MI and the diagnosis of LVPA varies from one day to 20 years (n=329); mean (95% CI) 5.1 (1.3, 8.9) months; according to this time interval, 29.0% of patients fell into the acute category (0, 14 days), 25.7% in subacute category (14, 90 days), and 45.5% into chronic category (more than 90 days) (Table 1).

Previous cardiac surgery was reported in 33.3% (95% CI:27.8, 39.2) of the cases (n=279), including CABG (21.1%) and valvular surgery (3.9%). History of smoking, hypertension, and diabetes mellitus was presented in 61.9% (24.2, 93.5), 52.2% (95% CI:21.8, 81.9), and 30.5% (CI 95%:16.9, 45.7) of patients, respectively. Previous cardiac surgery was reported in 33.3% (95% CI:27.8, 39.2) of the cases (n=279), including CABG (21.1%) and valvular surgery (3.9%). History of smoking, hypertension, and diabetes mellitus was presented in 61.9% (24.2 to 93.5), 52.2% (95% CI:21.8, 81.9), and 30.5% (CI 95%:16.9, 45.7) of patients, respectively (Table 1).

The ECG data on admission was available for 192 cases, of whom 185 (96.3%) had an abnormal finding(s), mainly ST-segment elevation (43.2% (95% CI:36.1, 50.1)), changes indicating old MI [42.7% (95% CI:35.6 to 50.0)], and/or tachycardia [9.4% (95% CI:5.6, 14.4)]. The location of infarction was, in order by frequency, inferior 37% (95% CI:21.8, 53.5), anterior 20.1 % (95% CI:11.6, 29.8), posterior 16.3% (95% CI:5.3, 31.0), and lateral in 9.0% (95% CI:5.9, 12.5).

The imaging findings are shown in table 3. Chest x-ray findings were reported in 92 patients; abnormalities cardiomegaly, cardiac mass, and cardiac silhouette sign were observed in [60.4% (95% CI: 50.2, 70.6), 35.1% (95% CI: 25.1, 45.1), and 25.2% (95% CI:16.1, 34.4)] of cases, respectively (Table 2).

**Table 2.**
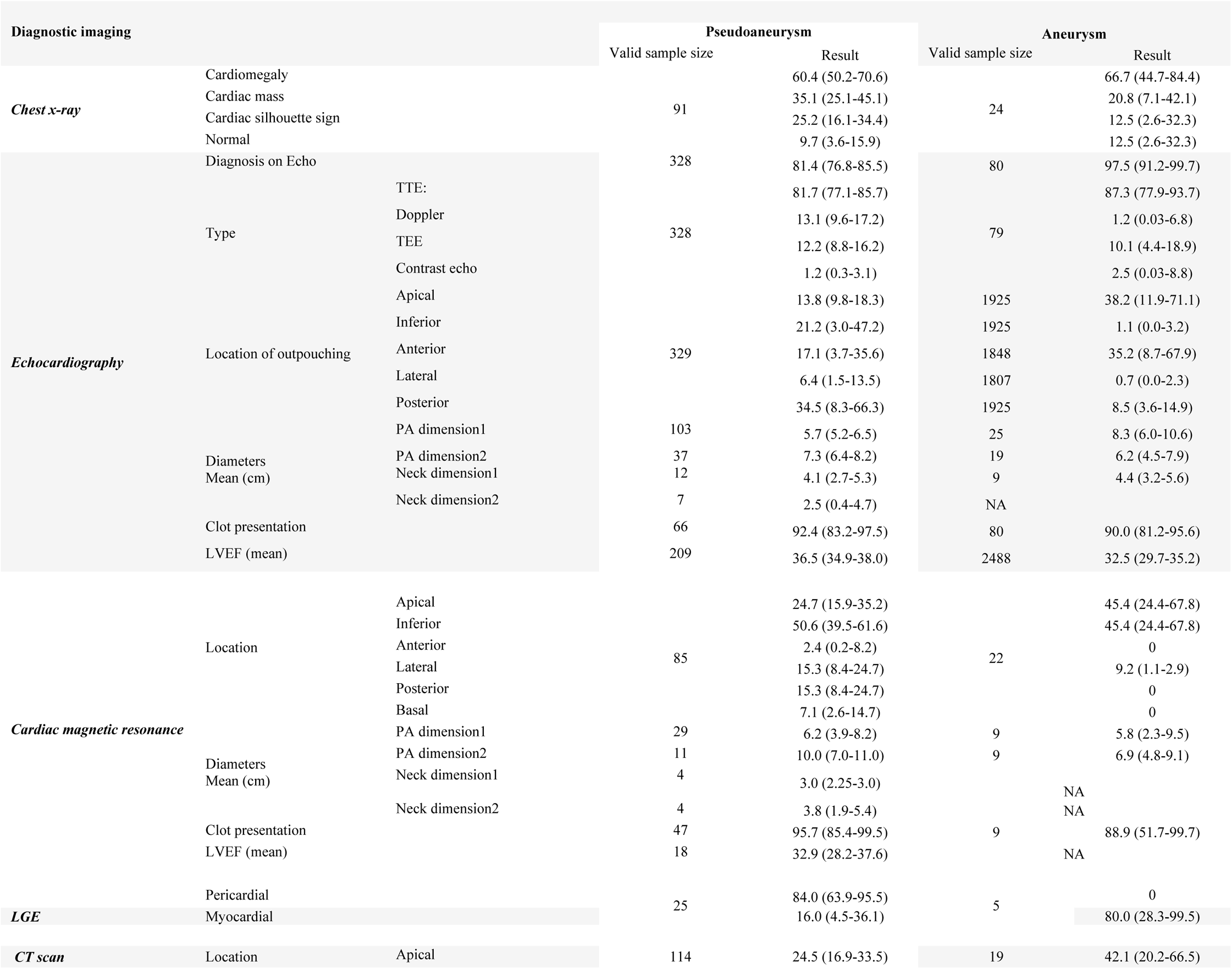
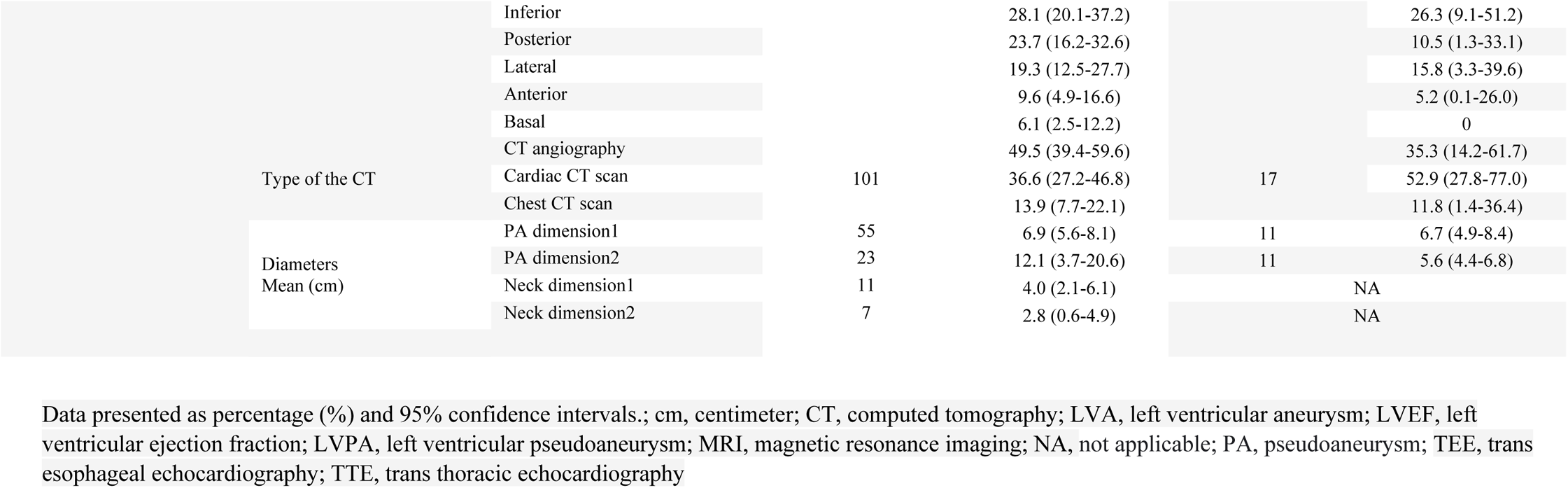
imaging findings of LVPA and LVA patients

Based on available data on Echocardiography evaluations (n=328), this imaging modality failed to diagnose LVPA in [18.6% (14.5, 23.2%)] of cases. The most frequent location of LVPA was posterior [34.5% (CI 95%:8.3, 66.3)], followed by inferior [21.2 (95% CI: 0.3, 47.2)], and anterior 17.1% (CI 95%:3.7, 35.6). The mean (95% CI) LVEF was 36.5% (34.9, 38.0) (n=209). The mean (95% CI) of the two reported dimensions of the LVPA were 5.7 (95% CI: 5.2, 6.5, n=103) and 7.3 (CI 95%: 6.4, 8.2) centimeters (Table 2).

Data on Cardiac magnetic resonance was available in 85 patients. Common locations of the LVPA were inferior at 50.6% (CI 95%: 39.5, 61.6). The mean (95% CI), LVEF on MRI, reported in 18 patients, was 32.9% (CI 95%: 28.2, 37.6). Late gadolinium enhancement was reported in 25 patients which majority of them had pericardial LGE [84.0% (CI 95%: 63.9, 95.5)] (Table 2).

A total of 114 cases underwent Computed tomography (CT) scan, mainly CT angiography (CTA) [49.5% (95% CI: 39.4, 59.6)]. The most prevalent location of LVPA in CT scans was inferior at 28.1% (95% CI: 20.1, 73.2), followed by apical at 24.5% (16.9, 33.5), posterior at 23.7% (95% CI: 16.2, 32.6). Hospital death and follow up death occur in 13.8% and 17.5% of LVPA cases, respectively (Table 2).

### Comparisons between LVPA and LVA

In both LVPA and LVA groups, over 70% of the cases were male (70.4% vs. 75.7 %, respectively), and their mean age fell in the seventh decade of life (65.4 vs. 60.8 years, respectively). Two groups were similar regarding the prevalence of cardiovascular disease risk factors, including; HTN, DM, hyperlipidemia, and smoking (Table 1).

On admission, while one out of three patients with LVA presented with arrythmia, only one out of 100 LVPA patients showed this symptom; however, cardiogenic shock was more prevalent among LVPA than LVA group (10.0% vs. 6.1%, respectively). In addition, patients with LVPA less frequently had dyspnea (43.3% Vs. 63.9 %, respectively) chest pain (39.2% Vs. 56.9%, respectively) and CHF symptoms (45.5% Vs. 59.7%, respectively) in comparison with those in LVA group (Table 1).

The mean time interval from MI to diagnosis was shorter in LVPA group than LVA (5.1 vs. 27.8, months, respectively). The most common previous cardiac surgery was CABG in both groups (Table 1).

On ECGs, LVPA group compared to LVA group more frequently showed ST-segment elevation (43.2% Vs. 28.6, respectively) but less frequently the changes indicating the old MI (42.2% Vs. 61.9, respectively). In addition, the most common location of infarction was inferior in the LVPA group [37% (95% CI:21.8, 53.5)] but the anterior segment in the LVA group [84.9% (95% CI: 66.8,97.0)] (Table 1).

On chest x-ray, a higher percentage of LVPA group had cardiac mass (35.1% vs. 20.8%) but a less percentage of them had evidence of cardiomegaly (60.4% vs. 66.7%) (Table 2).

Echocardiography had a lower sensitivity for diagnosis of LVPA compared to LVA group (81.4% vs. 97.5%). In contrast to LVPA that mainly located on posterior and inferior (34.5%,24.2%, respectively) segments, LVA involved apical and anterior segments (38.2%,35.2%, respectively). Patients with LVA had a higher mean (95% CI) LVEF than patients with LVPA [36.5% (34.9, 38.0) vs. 32.5% (29.7, 35.2)]. On cardiac MRI, approximately four out of five patients with LVPA (21/25) had a pericardial LGE, but none of five patients with LVA group had this pattern of LGE (Table 2).

The LVPA group compare to LVA group had a higher hospital mortality (13.8% vs. 4.7%, respectively) and follow up death (17.5% vs. 9.0%, respectively).

## Discussion

This comprehensive pooled clinical an imaging data of all available studies describing LVPA after MI and compared the findings with those in a pooled cohort of LVA patients.

Since 1991, a total of 504 cases of LVPA following MI have been reported worldwide. The reported cases predominantly were male, and their age varied from 33 to 97 years old. In About half of the cases, LVPA was diagnosed 90 days or more (chronic LVPA) after the last MI. CHF symptoms were reported in nearly one out of three cases at the presentation; also, cardiogenic shock occurred in 10% of them on admission. A little more than 20% of the patients had a previous history of CABG. The most common abnormalities on ECG were STE and changes indicating old MI, and on CXR was cardiomegaly and cardiac mass, respectively. Approximately 80% of LVPA were diagnosed by echocardiography; the most common locations of PA were posterior, followed by inferior.

The LVPA patients were roughly older than LVA patients but gender was mainly male in both groups. Regarding clinical symptoms on admission, arrhythmia was common in the LVA group but not in LVPA. LVPA patients had a shorter mean time interval from the last MI to diagnosis compared to LVA patients. Contrary to LVPA, the most common location of LVA was apical and anterior. LVEF was a little higher in the LVPA group. ST-segment elevation was observed less frequently in LVA cases, and cardiomegaly was the most common finding on CXR in these patients. Contrary to LVA, pericardial LGE was a common finding in the LVPA patients on cardiac MRI. Hospital mortality was about three times higher in the LVPA group compared to the LVA group. About 95% and 73% of LVPA and LVA cases were treated surgically, respectively.

Gender pattern in LVPA and LVA was roughly the same and similar to MI; about 70% of MI cases are male. However, LVA patients were slightly older than the LVPA group (473, 474).

Post-MI LVPA occurs when a pericardial adhesion restricts a true rupture of the ventricular wall, but LVA appears following cardiac remodeling that leads muscle fiber cells to change the geometry of the left ventricular cavity and form LVA (475). Aging is associated with the increased risk of impaired ventricular remodeling, a possible mechanism involved in LVA. In fact, elderly patients more frequently experience suppressed post-MI inflammation, which is vital to prevent ventricular remodeling (476).

A higher rate of arrhythmia observed in LVA than in LVPA could be justified by their different possible etiology; indeed, structural remodeling is associated with the development of areas of slow conduction and conduction block, resulting in the occurrence of arrhythmias (233, 477).

Also, LVPA patients may more likely develop cardiogenic shock than LVA patients due to a higher risk of cardiac tamponade and ventricular rupture. As LVPA occurs more commonly in the acute phase of the infarction but LVA in the chronic phase, LVPA was mainly characterized by STE and LVA by signs of old MI on ECG.

LVPA is a life-threatening and emergency condition and more frequently results in worse outcomes compared to LVA; based on our pooled analysis, LVPA patients dead approximately 2 to 3 times more than their counterparts in the LVA group during the hospitalization and after discharge.

Still, early diagnosis can lead to timely management and improve the prognosis in the affected patients. In this way, the distinction between different outpouchings of the left ventricle, i.e., LVPA and LVA, is imperative for choosing a proper treatment strategy. While almost all patients, with or without symptoms, in the LVPA group undergo cardiac surgery, management strategy in LVA is largely determined based on the presence of symptoms (478).

The availability, cost-effectiveness, and safety of echocardiography have made it the most utilized imaging modality in assessing structural heart diseases, including outpouchings of the left ventricle. Based on our pooled analysis, echocardiography has failed to detect the presence of LVAP in about 20% of cases but showed a better performance in diagnosing LVA (479). Some echocardiography features, such as the location of observed outpouchings and neck diameters, could also help to distinguish LVPA from LVA; contrary to LVA, LVPA is more frequently located on the posterior and inferior segments. Their different locations of infarction could explain this difference (480). However, another proposed explanation is that patients with anterior LVPA are at an increased risk of ventricular rupture leading to death before diagnosis (481).

Contrary to LVA, LVPA is usually characterized by a narrow neck on TTE echocardiography, which manifests as a turbulent flow on Doppler echocardiography (1,482); noteworthy, some chronic LVPA may have a wide base similar to LVA, causing misdiagnosis. However, most included studies failed to provide data on neck diameters in both groups.

MRI and CT scans, especially CT angiography, have higher accuracy than echocardiography for diagnosis of cardiac outpouchings, including LVPA; though, in practice, their utilization is limited by factors such as less availability, no portability, and high costs (483).

Available evidence suggests that pericardial LGE can be a helpful tool in differentiating LVPA from LVA [2]. Indeed, LGE presents in the wall of the aneurysm sac in both LVPA and LVA on cardiac MRI, but delayed enhancement of the pericardium is thought to be a characteristic feature of pseudoaneurysm. It indicates the presence of pericardial inflammation and fibrosis induced by leakage of blood into the pericardial space subsequent to ventricular rupture (484, 485).

### Limitations and strengths

As a review of published studies, our pooled data sets were incomplete for some variables, especially imaging features, likely due to the retrospective nature of the design of the included studies. Also, our review lacked confirmation of imaging findings by a core laboratory as we pooled data from published case series /reports. However, given the detailed protocols reported by most studies regarding the actual images, the validity of imaging findings can be considered acceptable.

using a comprehensive search of online databases, we included all relevant studies describing patients with post-MI LVPA; Given the small number of patients reported by each case series/case reports, this pooled cohort can promote the current understanding of the clinical and imaging findings in LVPA after MI. we also and compared the findings in LVPA with those in a pooled and large cohort of LVA patients to help physicians for differentiating these outpouchings.

## Conclusion

Our pooled cohort suggests that LVPA is probably located on the posterior and inferior segments in most patients, contrary to the LVA that mainly involved the anterior and apical segments. Arrhythmia is likely a common clinical finding in LVA but not LVPA. Most ECG abnormalities might be ST-Elevation in LVAP and changes suggesting old MI in LVA. Cardiac mass on CXR may represent the presence of LVPA more than LVA. Echocardiography may fail to detect the presence of LVPA in one out of five patients. On cardiac MRI, pericardial LGE may suggest the presence of LVPA than LVA in suspected patients.

## Data Availability

Not applicable.

## Acknowledgment

Researchers appreciated the Clinical Research Development Units of Kamali and Rajaee Hospitals in Alborz University of Medical Sciences.

